# Fluvastatin mitigates SARS-CoV-2 infection in human lung cells

**DOI:** 10.1101/2020.07.13.20152272

**Authors:** Francisco J. Zapatero-Belinchón, Rebecca Moeller, Lisa Lasswitz, Marco van Ham, Miriam Becker, Graham Brogden, Ebba Rosendal, Wenjie Bi, Belén Carriquí, Koushikul Islam, Annasara Lenman, Antonia P. Gunesch, Jared Kirui, Thomas Pietschmann, Anna K Överby, Lothar Jänsch, Gisa Gerold

**Author notes:** equally contributed (Francisco J. Zapatero-Belinchón, Rebecca Moeller, Lisa Lasswitz).

## Abstract

Clinical data of patients suffering from COVID-19 have indicated that statin therapy, used to treat hypercholesterolemia, is associated with a better clinical outcome. We therefore investigated the effect of statins on SARS-CoV-2 infection in human lung cells and found that fluvastatin inhibited coronavirus infection, while other tested statins did not. Fluvastatin inhibited high and low pathogenic coronaviruses *in vitro* and *ex vivo* in a dose-dependent manner. Proteomic analyses of infected versus uninfected lung epithelial cells treated with fluvastatin, simvastatin, or rosuvastatin revealed that all tested statins modulated the cholesterol synthesis pathways without compromising the innate antiviral immune response. Strikingly, fluvastatin treatment uniquely affected the proteome of SARS-CoV-2 infected cells, specifically downregulating proteins that modulate protein translation and viral replication. These results suggest that statin therapy poses no additional risk to individuals exposed to SARS-CoV-2 and that fluvastatin may have a moderate beneficial effect on SARS-CoV-2 infection by modulating protein translation.

## Introduction

SARS-coronavirus 2 (SARS-CoV-2), the causative agent of coronavirus disease 2019 (COVID-19), has infected millions of people worldwide^1^. There are inter-individual differences in the course of COVID-19, which are incompletely understood to date^2^. The SARS-CoV-2 receptor ACE2^3^ is localized in lipid rafts, which are cholesterol and glycosphingolipid enriched microdomains of the plasma membrane^4,5^. Consequently, SARS-CoV, which also uses ACE2 as host receptor, and other pathogens require such lipid rafts at different life cycle steps^6–9^. Statins are cholesterol-lowering drugs, prescribed to treat hypercholesterolemia, cardiovascular disease and diabetes. Notably, a significant proportion of the population takes statins, in particular the elderly^10^, who are at high risk of developing severe symptoms upon SARS-CoV-2 exposure^11^. Retrospective studies report a correlation of statin therapy and reduced all-cause mortality of COVID-19 patients^12,13,^. On the other hand, statins may increase expression of the ACE2 receptor of SARS-CoV-2^14,15^. Whether statins not only influence disease outcome but also directly impact SARS-CoV-2 replication is unknown.

In this study, we used coronavirus cell culture models to assess a possible direct impact of statins on coronavirus infection. First, we investigated the role of selected statins in low pathogenic coronavirus 229E infection. Then, we validated our findings with SARS-CoV-2 using cell lines and air-liquid interface cultures from differentiated human primary bronchial epithelial cells. To determine the effects of statins on cholesterol biosynthesis and possible secondary effects influencing SARS-CoV-2 infection, we analyzed the cellular proteome in the presence or absence of statins and SARS-CoV-2. Statin treatment upregulated cholesterol synthesis enzymes in infected and uninfected cells without impairing viral immune sensing. Interestingly, fluvastatin treatment uniquely affected the proteome of SARS-CoV-2 infected cells, specifically downregulating pathways and proteins important for RNA degradation, protein translation, and viral replication. Altogether, our results indicate that statin treatment is safe during the COVID-19 pandemic and that fluvastatin may inhibit SARS-CoV-2 infection by modulation of viral replication and translation.

## Results

### Selected statins reduce coronavirus infection in human cells

To assess a possible direct impact of statins on coronavirus infection, we infected Huh7.5 cells expressing a Firefly luciferase (Huh7.5 Fluc) with the low pathogenic human coronavirus 229E. At a concentration of 5 μM statins reduced infection of CoV-229E moderately between 10 – 40 %. This reduction of susceptibility was most pronounced (40%) and statistically significant for fluvastatin (Fig. 1a). The most commonly prescribed statins simvastatin and atorvastatin reduced coronavirus 229E infection up to 10%. Lovastatin, pravastatin and rosuvastatin reduced coronavirus 229E infection up to 15%, 25%, and 30%, respectively, but the observations did not reach statistical significance in the cell culture assay. Chloroquine completely blocked coronavirus 229E infection as expected^16,17^. When performing a dose-response analysis for fluvastatin, we found a dose-dependent reduction of coronavirus infection with a half-maximal inhibitory concentration of 9.8 μM. At the investigated fluvastatin doses, cell viability remained at 80% and higher indicating no major cytotoxic effects (Fig. 1b). Dose-response analysis for rosuvastatin showed a 40% reduction of coronavirus 229E infection at 50 µM, which correlated with a decrease in cell viability (Fig. 1c). We did not observe a dose-dependent reduction of coronavirus 229E infection upon simvastatin treatment (Fig. 1d).

**Fig. 1:**
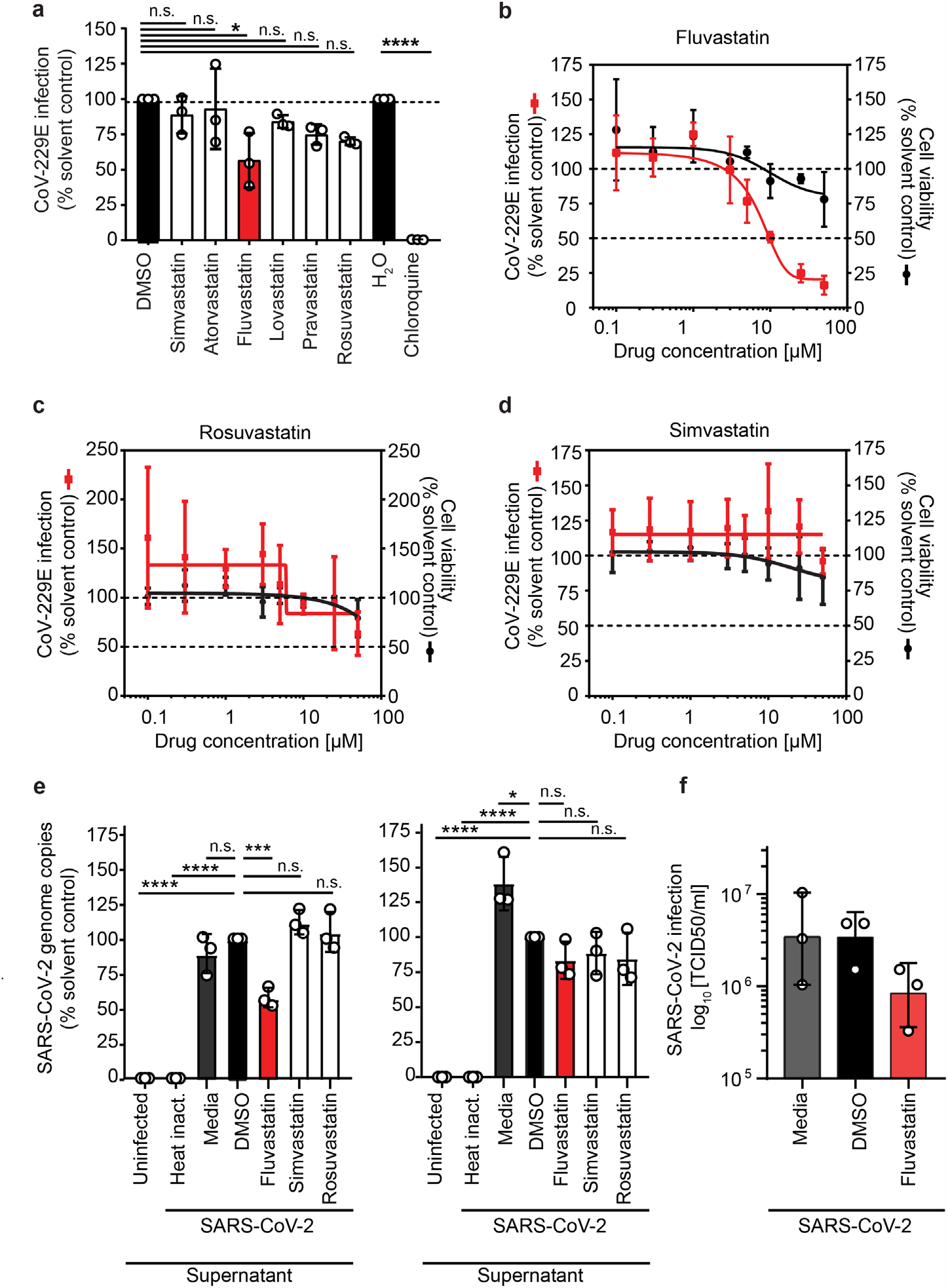
Lipid-lowering drugs selectively reduce coronavirus infection in human cells. **a**, Huh7.5 Fluc cells were pretreated 24 h with the indicated statins (5 μM) or chloroquine as positive control and then infected with hCoV229E harboring a Renilla reporter gene (MOI 0.005) in presence of the drugs. Renilla luciferase activity as measure of infectivity was determined 48 h after infection. The hCoV229E infectivity (Renilla luciferase activity) was normalized to cell viability measured by the constitutive expression of Firefly luciferase in Huh7.5 Fluc cells. The dotted lines indicate 50% and 100% of infection in solvent control treated cells. **b-d**, Dose-response curves for the antiviral and cytotoxic effect of fluvastatin (b), rosuvastatin (c) and simvastatin (d) in human cells. Infection and statin treatment was performed as in a. at the indicated statin concentration. **e**, Fluvastatin decreases SARS-CoV-2 susceptibility of human respiratory epithelial Calu-3 cells. Virus genome copy numbers in cell lysates and supernatants from cells pretreated with selected statins (10 μM) or DMSO solvent control and infected with SARS-CoV-2 (MOI 2.0×10^−5^) are shown. Results are normalized to viral copy numbers in DMSO treated cells. **f**, Number of infectious SARS-CoV-2 particles in cell culture supernatant from fluvastatin pretreated and SARS-CoV-2 infected Calu-3 cells (as in e) was determined by titration on Vero cells. (a-e) Mean ± SD of three independent biological replicates shown. One-way ANOVA, followed by Dunnett’s multiple comparison test * p<0.05, *** p<0.0005, **** p<0.0001.

To investigate whether statins could modulate infection with highly pathogenic SARS-CoV-2 isolates, we pretreated human respiratory epithelial cells with 10 µM fluvastatin, rosuvastatin or simvastatin and infected the cells with SARS-CoV-2 (strain SARS-CoV-2/München-1.2/2020/984,p3)^18^. SARS-CoV-2 genome copy numbers significantly decreased down to 60% in supernatants upon fluvastatin treatment, whereas rosuvastatin or simvastatin treatment did not reduce SARS-CoV-2 genome copy numbers in supernatants (Fig. 1e). In cell lysates of infected cells, treated with fluvastatin, rosuvastatin or simvastatin, a slight reduction of about 15% of SARS-CoV-2 genome copy numbers was detected, but did not reach statistical significance (Fig. 1e). As we detected a significant decrease in SARS-CoV-2 genome copy numbers in supernatants upon fluvastatin treatment, we titrated released infectious SARS-CoV-2. Fluvastatin treatment reduced SARS-CoV-2 titers 4-fold compared to DMSO treatment, as determined by TCID50 assay on Vero cells (Fig. 1f). Collectively, our data highlight that statins do not promote coronavirus infection in human cells as was speculated, but instead selected statins attenuate coronavirus infection *in vitro*.

### Fluvastatin moderately decreases SARS-CoV-2 infection in air-liquid interface cultures of human primary bronchial epithelial cells

We next aimed to corroborate our findings for fluvastatin treatment in a model that closely resembles the human respiratory tract. To that end, we differentiated human primary bronchial epithelial cells (HBECs) isolated from three different individuals at an air-liquid interface (ALI) into a pseudostratified, polarized epithelium containing an apical layer of fully functional secretory and ciliated cells and an underlying layer of basal cells. SARS-CoV-2 infection was assessed in these ALI cultures with or without fluvastatin (10 µM or 50 µM) pretreatment administered in the basolateral medium. Here, we increased the dosage of fluvastatin above the IC50 determined in monolayer cell culture, as in this more complex multilayer cell culture model higher doses might be required.

To monitor the course of infection, we measured progeny virus release into the apical mucus with real time q-PCR in 24 h intervals. Viral RNA pronouncedly increased over 72 h (10^5^ to 10^7^ for donor #1 and #2; 10^4^ to 10^6^ for donor #3; Fig. 2a black lines) indicating active viral replication in the ALI cultures. We detected varying levels of viral RNA in the samples from different donors (donor #1 and #2 vs. donor #3), which may reflect differences in susceptibility of individuals as observed in natural infection. Treatment with 10 µM fluvastatin did not affect viral release in highly infected samples (donor #1 and #2; Fig. 2a, red dashed lines), whereas a moderate decrease in released virus was observed in a less infected sample (to 10^5^ in donor #3; Fig. 2a, red dashed line). In contrast, treatment with 50 µM fluvastatin, dramatically reduced viral release in samples from all donors (at least two log steps, Fig. 2a, dark red dotted lines).

We next stained the HBEC ALI cultures for differentiation markers of ciliated cells (acetylated tubulin, Fig. 2c, yellow) and goblet cells (mucin5AC, Fig. 2c, red) in addition to SARS-CoV-2 nucleocapsid protein (SARS-CoV-2 NC) (Fig. 2c, green), and acquired overview images of all samples, as well as detailed confocal images of highly infected HBEC ALI cultures. We observed differences in number of goblet and ciliated cells between the donors, which may reflect donor-dependent variation or slight differences in the differentiation. SARS-CoV-2 infection was mainly detected in ciliated cells as reported previously for ALI cultures^19^, but also in unstained cell populations, that most possibly represent basal cells or differentiation intermediates as reported for human lung tissues^20^.

**Figure 2:**
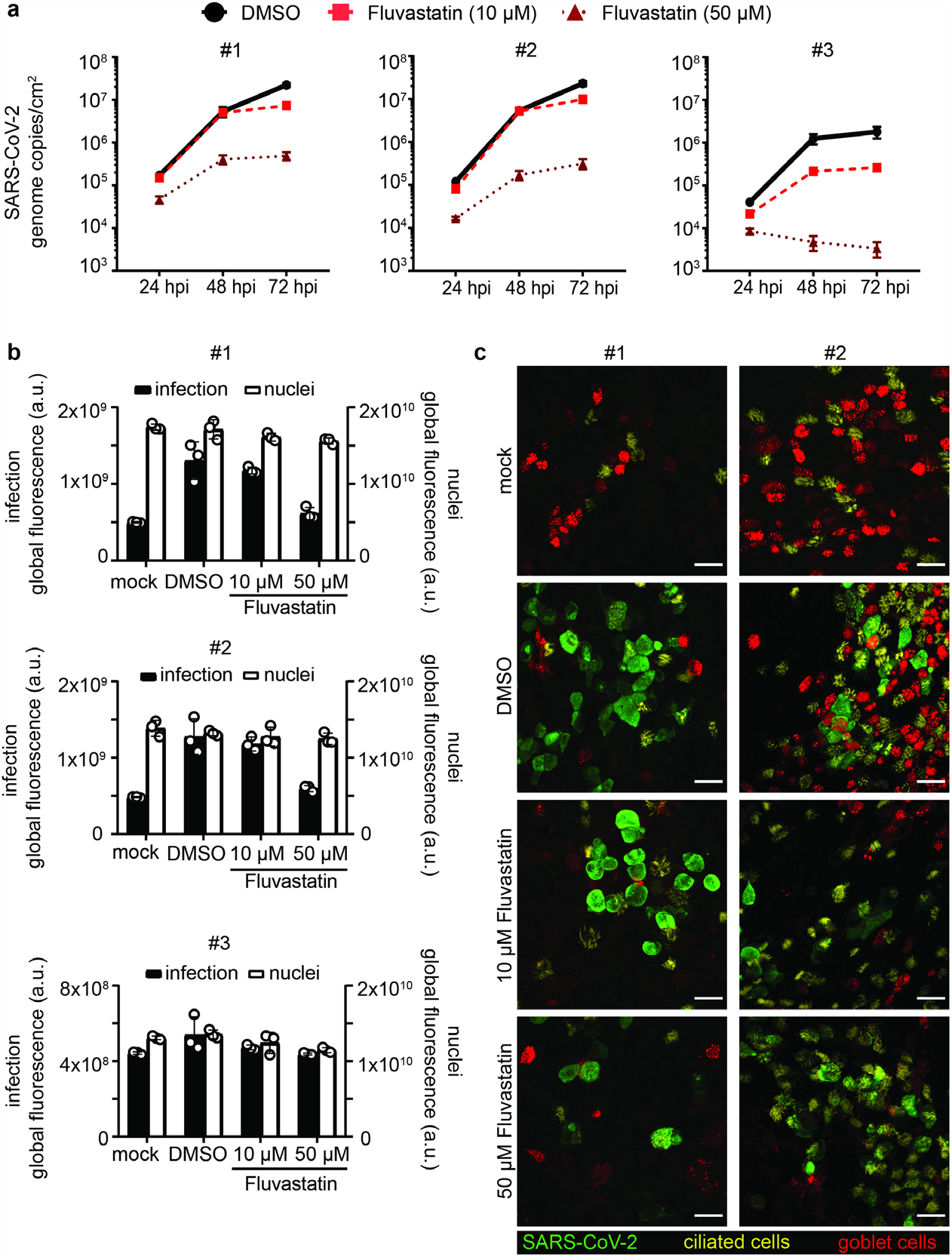
Fluvastatin treatment reduces SARS-CoV-2 infection in HBEC ALI cultures. **a-c**, HBEC ALI cultures from three donors were pretreated with DMSO solvent control or fluvastatin (10 µM or 50 µM) in the basal media for 24 h prior to SARS-CoV-2 infection at the apical side (4.5×10^4^ PFU). To assess viral replication, apical samples from HBEC ALI cultures were collected 24, 48 and 72 h post infection by a 1 h wash in 300 µL growth medium. Progeny virus released within these 24 h periods were quantified by RT q-PCR alongside an RNA standard. RNA genomes per ALI area in DMSO-treated samples (circle, black solid line) was plotted against time for the different statin concentrations (10 µM: square, red dashed line; 50 µM: triangle, dark red dotted line). RT q-PCRs were run in duplicates from three HBEC ALI inserts per sample. **b**, ALI cultures were fixed at 72 h (endpoint) and stained for SARS-CoV-2 nucleocapsid protein and nuclei and analyzed by immunofluorescence. Global fluorescence intensity of infection (black bars) and nuclei (white bars) was quantified from a fixed ROI omitting the well borders. Depicted are averages and SD in global fluorescence in arbitrary units from triplicates per condition from HBEC ALIs for each donor. **c**, ALI samples at 72 h (endpoint) were stained for SARS-CoV-2 nucleocapsid protein for infection (green), mucin5AC for secretory cells (red) and acetylated tubulin for ciliated cells (yellow) and imaged on a scanning confocal microscope. Depicted are maximum intensity projections from z-stacks of representative sites for the different conditions from two different individuals. Scale bars = 25 µm.

To validate our findings, we quantified the intracellular infection levels from immunofluorescence staining for SARS-CoV-2 NC on the HBEC ALI cultures at 72 h. Sample integrity was assessed at the same time by nuclear staining. We observed that the nuclear signal remained constant upon infection or fluvastatin treatment for all samples (Fig. 2b, white bars), indicating no cellular loss upon treatment and infection. SARS-CoV-2 NC positive cells appeared at many sites in the cultures for the highly infected samples from donor #1 and donor #2, whereas the samples from donor #3 showed only a few sites with SARS-CoV-2 NC positive cells (Fig. 2b, black bars, DMSO and Fig. S1), in line with the data on virus release. Fluvastatin treatment reduced the overall SARS-CoV-2 NC signal only slightly at lower dose (Fig. 2b, black bars, 10 µM and Fig. S1) and highly drastic at the higher dose (Fig. 2b, black bars, 50 µM and Fig. S1).

In summary, these results suggest that fluvastatin treatment does not enhance SARS-CoV-2 infection in three-dimensional respiratory epithelium models but may rather have a moderate attenuating effect similar as seen in the monolayer cell culture system.

### Statins regulate cholesterol synthesis independently of SARS-CoV-2 infection

To determine the effects of fluvastatin, rosuvastatin and simvastatin on global cellular processes, amongst which cholesterol biosynthesis, Calu-3 cells were pretreated with each statin or solvent control, then infected with SARS-CoV-2 or left uninfected (mock), and cellular proteomes were analyzed by label free mass spectrometry (Fig. 3a). As additional control of inoculum induced changes in the absence of replicating virus, Calu-3 cells incubated with heat-inactivated virus were used. All experiments were performed in biological triplicates, generating quantitative information of in total 27 proteomes. SARS-CoV-2 genome copies decreased significantly in supernatants of fluvastatin treated cells compared to DMSO treated cells, in line with our previous experiments (Fig. S2a). In total 3229 proteins were identified unambiguously by at least one unique tryptic peptide. Global inspection of our proteomic data revealed four cholesterol biosynthesis pathway enzymes that were significantly more abundant (either - log_10_(p-value) ≥ 2, or log_2_(fold change) ± 4 and - log_10_(p-value) ≥ 1.3) upon treatment with fluvastatin, namely 3-hydroxy-3-methylglutaryl-CoA reductase (HMGCR), farnesyl-diphosphate farnesyltransferase 1 (FDFT1), squalene monooxygenase (SQLE) and methylsterol monooxygenase 1 (MSMO1) (Fig. 3b, shown in orange). In addition, expression levels of twelve proteins unrelated to cholesterol biosynthesis changed significantly (shown in blue). To investigate the modulatory effects of each statin during SARS-CoV-2 infection in more detail, we compared the top 40 altered cellular pathways for each experimental condition. All three statins upregulated cholesterol biosynthesis pathways and depleted Ran, melatonin, and androgen signaling pathways (Fig. 3c). The upregulation of cholesterol biosynthesis enzymes to antagonize statin-mediated blockade of HMGCR is in line with previous reports^21^. Expectedly, statin treatment led to a slight decrease in cholesterol concentration in cells cultured in the absence of FCS, whereas no decrease in cholesterol was observed in cells cultured in medium containing FCS, as used in our experimental conditions (Fig. S2b). Importantly, cholesterol synthesis proteins were upregulated in both the presence and absence of SARS-CoV-2 infection (Fig. 3d). Specifically, HMGCR, SQLE and FDFT1 expression levels increased upon treatement with fluvastatin, simvastatin and rosuvastatin independently of SARS-CoV-2 infection (Fig. 3e). This strongly suggests that SARS-CoV-2 infection itself does not impair statin-mediated cholesterol synthesis inhibition (Fig. 3d and e).

**Fig. 3:**
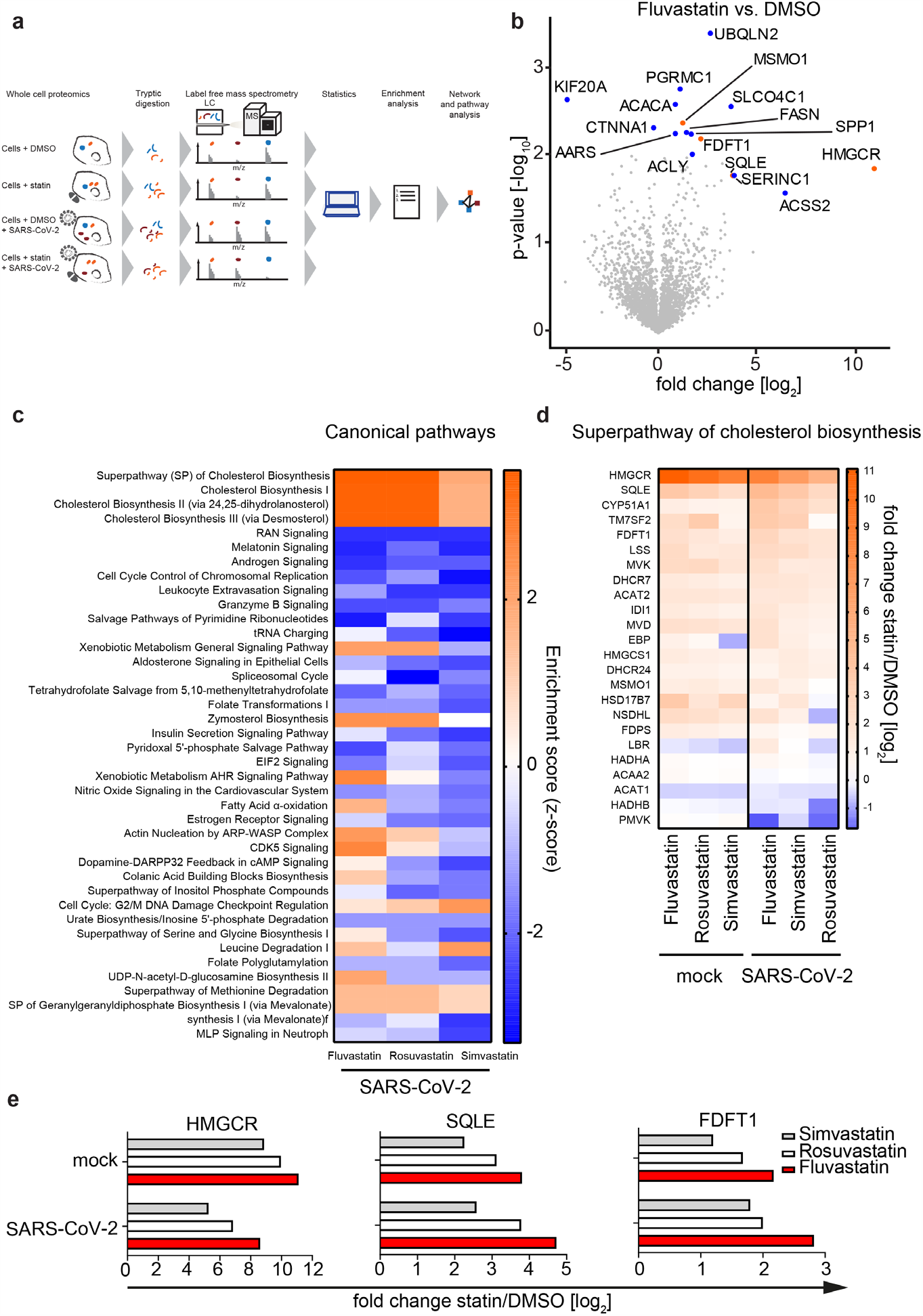
Statin treatment upregulates expression of proteins involved in cholesterol synthesis. **a**, Experimental workflow of the whole cell shotgun proteomics analysis. Three independent passages of Calu-3 cells were pretreated with 10 μM of the different statins or solvent control for 24 h and subsequently infected in the presence of the drug with SARS-CoV-2 (MOI 2.0×10^−5^) or left uninfected (mock). 48 h later, viral titer was determined via RT-qPCR from the supernatants (see Fig. S2a) and cells were lysed and used for protein extraction and subsequent analysis by label-free mass spectrometry. Unambiguous tryptic peptides were identified, protein abundance determined by label-free quantification algorithms and compared between different experimental conditions. **b**, Volcano plot showing changes in protein abundance in fluvastatin treated cells vs. DMSO solvent control treated cells. Significantly enriched proteins (either –log10 p-value ≥ 2 or log2 fold change ≥ ±4 with – log10 p-value ≥ 1.3) are highlighted in blue, significantly enriched proteins involved in the cholesterol synthesis pathway are highlighted in orange. n=3, significance was analysed using the Student’s t test. **c**, Analysis of top 40 significantly enriched canonical pathways (p-value ≤ 0.05) upon statin treatment in SARS-CoV-2 infected Calu-3 cells. **d**, Statin induced changes in protein abundance in the superpathway of cholesterol synthesis in SARS-CoV-2 infected cells and uninfected cells (mock). **e**, Median intensity differences from three independent experiments of selected cholesterol synthesis pathway proteins upon indicated statin treatment. Experiments performed in biological triplicates.

### Statin treatment does not impair the innate immune response against SARS-CoV-2

During SARS-CoV-2 infection, the innate immune response plays an important role to fight the virus^22^. However, immediate immune response is also causing the observed immunopathology^23^. Therefore, it is crucial to assess how prescribed medications, like statins, modulate host defense mechanisms in the context of SARS-CoV-2 infection. To demonstrate the effect of SARS-CoV-2 infection in statin treated cells, we compared protein expression in fluvastatin treated uninfected cells with fluvastatin treated and SARS-CoV-2 infected cells (Fig. 4a). As expected, we found viral proteins such as the spike protein (S), the matrix protein (M) as well as the accessory protein 9b (Orf9b) in lysates from infected cells in all conditions (red dots in Fig. 4a for fluvastatin treated conditions). In addition, virus infection in fluvastatin treated cells upregulated host proteins such as the interferon-induced GTP-binding protein Mx1 (MX1), transcription termination factor 2 (TTF2), WD repeat-containing protein 87 (WDR87) and DENN domain containing 6A (DENND6A). Among the downregulated proteins upon infection were host proteins such as septin-8 (SEPT8) and nuclear receptor-binding protein (NRBP1). We next analyzed, which upstream regulators accounted for the observed differences in protein abundance in uninfected and infected cells in presence or absence of statins (Fig. 4b). Here, we additionally included the protein abundance profile of cells incubated with a heat-inactivated virus control to account for protein dysregulation caused by the inoculum in the absence of viral genome replication. Interestingly, we observed that tumor necrosis factor (TNF)-modulated proteins were strongly downregulated in infected cells treated with DMSO, simvastatin or rosuvastatin, while fluvastatin treatment prevented this downregulation. In fact, SARS-CoV-2 infected cells pretreated with fluvastatin displayed similar TNF-modulated protein abundance patterns as cells inoculated with the heat-inactivated virus control. In contrast to the downmodulating effect on the TNF pathway, SARS-CoV-2 infection increased the expression of proteins regulated by interferon lambda-1 (IFNL1) and interferon alpha-2 (IFNA2). This IFN pathway induction occurred both in the absence and in the presence of all three statins (enrichment scores between 2 and 3), while it was absent in cells incubated with heat-inactivated virus control. This suggests that sensing of the virus in our cell culture system depends on active viral replication and that this sensing is not impaired by statins. Next, we examined the protein abundance of genes regulated by TNF, IFNL1, and IFNA2 in uninfected and infected cells (Fig 4c-d). SARS-CoV-2 infection downregulated TNF-dependent proteins such as heparan sulfate proteoglycan 2 (HSPG2) or syndecan 4 (SDC4) independent of statin treatment (Fig. 4c). In contrast, virus infection strongly upregulated the interferon induced protein with tetratricopeptide repeats 3 (IFIT3) and IFIT1 only in DMSO, simvastatin and rosuvastatin treated cells, but not fluvastatin treated cells. Indeed, fluvastatin treated SARS-CoV-2 infected cells showed protein expression patterns of intermediate phenotype in-between the phenotypes of heat-inactivated virus incubated cells and DMSO treated SARS-CoV-2 infected cells. The interferon-inducible protein MX1 was downregulated in statin-treated uninfected cells compared to DMSO treated uninfected cells, but was highly induced upon SARS-CoV-2 infection (Fig. 4d and e). Interestingly, we found the interferon-inducible proteins 2’-5’-oligoadenylate synthetase 3 (OAS3), ISG15 ubiquitin like modifier (ISG15), DExD/H-box helicase 58 (DDX58), and major histocompatibility complex, class I, C (HLA-C) to be upregulated by statin treatment in uninfected cells. This suggests a possible modulatory role of statins on the innate immune response of the respiratory epithelium (Fig. 4d and e). In sum, statins did not strongly affect the innate immune response to SARS-CoV-2, but fluvastatin slightly decreased innate immune responses to SARS-CoV-2 infection as compared to DMSO, simvastatin and rosuvastatin treatment.

**Fig. 4:**
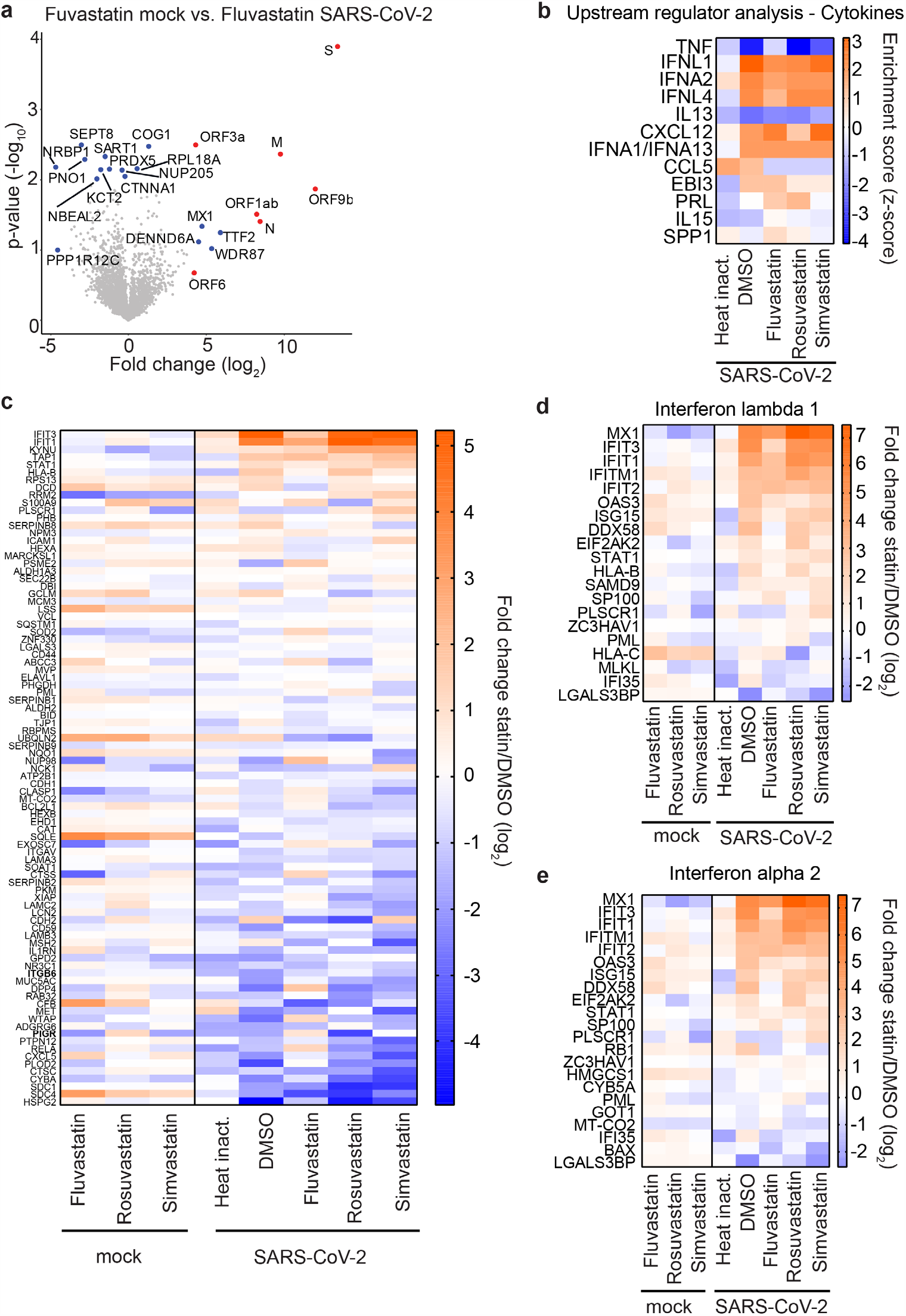
Statin treatment does not alter the innate immune response to SARS-CoV-2. **a**, Volcano plot showing changes in cellular protein abundance upon SARS-CoV-2 infection of fluvastatin treated cells. Viral proteins are depicted in red and significantly modulated host proteins in blue (either –log10 p-value ≥2 or log2 fold change ≥ ±4 with –log10 p-value ≥1.3). n=3, significance was analysed using the Student’s t test. **b**, Heatmap showing upstream regulatory cytokines and chemokines affecting protein abundance in cells upon SARS-CoV-2 infection and statin treatment (red upregulated pathway; blue: downregulated pathway). **c-e**, Changes in TNF regulated (c) and interferon stimulated genes (d,e) upon SARS-CoV-2 infection and statin treatment. Heatmap representing the log2 fold change in protein abundance upon statin treatment as compared to the DMSO solvent control in uninfected (mock) or SARS-CoV-2 infected cells. Heat inact.; Replication deficient heat inactivated virus control. Median intensities from three biological replicates.

### Fluvastatin induces a unique protein expression profile in SARS-CoV-2 infected cells compared to simvastatin and rosuvastatin

Our results suggest that fluvastatin may inhibit infection independently of its main cellular target HMGCR (Fig. 3). To elucidate secondary fluvastatin-specific effects on SARS-CoV-2 infected cells, we compared protein expression levels in infected cells with and without statin treatment. To focus on highly regulated proteins, we included proteins with either a -log_10_(p-value) ≥ 2 or with a log_2_(fold change) ± 4 and a -log_10_(p-value) ≥ 1.3 in our analysis (Fig. 5a). Eleven proteins were upregulated in infected cells treated with fluvastatin as compared to infected cells treated with DMSO. Expectedly, HGMCR was upregulated strongest upon fluvastatin treatment with a 380-fold increase over DMSO infected cells. Seven proteins, including the E3 ubiquitin ligase cullin CUL4B and the cytoskeletal forming GTPase SEPT8, were downregulated upon fluvastatin treatment. In addition, we detected seven viral proteins of which fluvastatin pretreatment reduced their expression, with N and ORF3a protein abundance being significantly reduced (Fig. S3). This is in line with the observed reduced virus infection measured by RT-qPCR and plaque assay (Fig. 1 and Fig. S2a). Next, we unbiasedly clustered protein expression levels of statin or DMSO-treated infected cells according to the differentially regulated pathways in each dataset (Fig. 5b and Fig. S4). Strikingly, we found that fluvastatin treated infected cells clustered independently from simvastatin, rosuvastatin or DMSO treated infected cells. This confirms that fluvastatin differentially affects the cellular proteome of SARS-CoV-2 infected cells. The cluster of pathways exclusively downregulated by fluvastatin included inhibition of ARE-mediated mRNA degradation, pyrimidine ribonucleotide de novo biosynthesis, telomerase signaling, PI3K/AKT signaling, protein kinase A signaling, and the unfolded protein response (Fig. S4). Among the pathways exclusively upregulated by fluvastatin, we found systematic lupus erythematosus in T cell signaling, UDP-N-acetyl-D-galactosamine biosynthesis II, CDK5 signaling, and the spliceosomal cycle (Fig. S4). These results suggest that fluvastatin treated infected cells may display stronger mRNA degradation and spliceosomal turnover than solvent, simvastatin, or rosuvastatin treated infected cells.

**Fig. 5:**
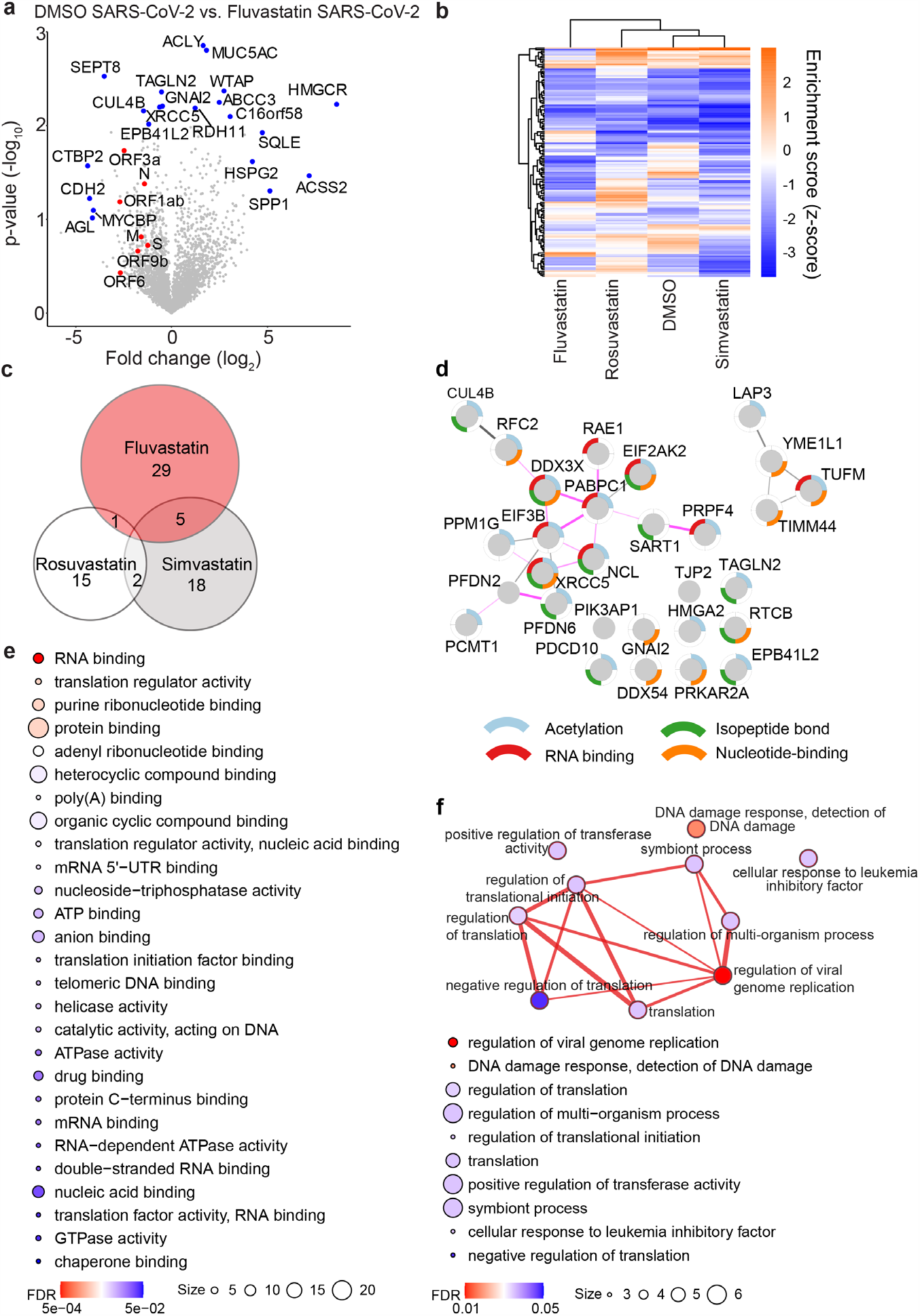
Fluvastatin induces a unique translational profile during SARS-CoV-2 infection. **a**, Volcano plot showing fluvastatin induced changes in cellular protein abundance in SARS-CoV-2 infected cells. Viral proteins shown in red and significantly modulated host proteins in blue (either –log10 p-value ≥ 2 or log2 fold change ≥ ±4 with –log10 p-value ≥ 1.3). n=3, significance was analysed using the Student’s t test. **b**, Canonical pathway analysis of whole cell proteome from SARS-CoV-2 infected cells with indicated treatments with a -log10 p value of at least 2. Hierarchical clustering based on enrichment scores. For a magnified heatmap including pathway information please refer to Fig. S4. **c**, Venn diagram of proteins significantly downregulated in SARS-CoV-2 infected cells upon treatement with the indicated statins. Numbers indicate statin specific and overlapping proteins for the indicated treatment. **d**, STRING network of proteins downregulated during SARS-CoV-2 infection in a fluvastatin specific manner. Nodes represent proteins and lines predicted interactions with line thickness indicating the strength of evidence (pink lines: experimental evidence; grey lines: co-expression, textmining, co-occurrence, database). Node border colors indicate the top four enriched gene ontology terms as indicated. **e**, Enriched gene ontology functions from fluvastatin downregulated proteins. Circle size signifies number of proteins with the indicated function and color calculated FDR value. **f**, Top, map depicting the enriched gene ontology biological processes within the dataset of fluvastatin downregulated proteins. Nodes represent biological process term and line thickness level of gene overlap between processes. Network node color displays FDR values. Bottom, list of enriched biological process terms indicating number of proteins per term (size) and FDR values (color).

To shed more light on the proteins specifically downregulated (p-values < 0.05) by fluvastatin, but not simvastatin and rosuvastatin in SARS-CoV-2 infected cells, we analyzed the differential abundance of proteins in infected cells treated with each statin in comparison to DMSO. Fluvastatin decreased the abundance of 35 proteins, while simvastatin and rosuvastatin decreased the abundance of 25 and 18 proteins in infected cells. Strikingly, we did not find any protein downregulated by all three statins. In addition, there was little overlap between pairs of two statins (Fig. 5c), suggesting that statins differentially downregulate protein expression upon infection. In total, 29 of the 35 proteins downregulated in their abundance by fluvastatin, were specific to this statin. To determine whether these 29 proteins are interconnected, we performed a STRING network analysis. We found two major clusters of interacting proteins in our dataset that were significantly enriched (PPI enrichment: 3.96×10^−4^) (Fig. 5d). The first cluster comprised four proteins and contained a mitochondrial protein involved in mitochondrial matrix peptide translocation (TIMM44) and a regulator of innate RNA recognition (TUFM). The second cluster comprised 15 proteins, most of which can be acetylated (12/15), have an isopeptide bond (10/15), or bind RNA (8/15) (compare color-coding of the node border). Moreover, according to experimental evidence most proteins in the second cluster interact with each other (13/15) (compare pink edges). Among these proteins we found the mRNA poly(A) tail binding protein polyadenylate-binding protein 1 (PAPBC1), the eukaryotic initiation factor 3 subunit B (EIF3B), the RNA helicase DEAD-Box helicase 3 X-Linked (DDX3X), and the IFN-induced protein kinase R (EIF2AK2, or PKR). Gene ontology analysis of the associated molecular functions of the downregulated proteins revealed, amongst others, enrichment of proteins involved in RNA binding (FDR = 5.4×10^−4^), translation regulator activity (0.0026), poly(A) binding (0.0057), and mRNA 5’-UTR binding (0.0074) (Fig. 5e). This suggests that the proteins specifically downregulated in fluvastatin treated SARS-CoV-2 infected cells play a role in translational regulation. When we compared the biological processes associated with all 29 proteins downregulated by fluvastatin in infected cells, we observed that the enriched biological processes are tightly interconnected (Fig. 5f). Whereas most of the enriched biological processes were related to protein translation, the most significantly enriched gene cluster was annotated as regulation of viral genome replication (FDR = 0.0103) that contained EIF2AK2, PABPC1, DDX3X, and high mobility group box 2 (HMGA2). In summary, these findings suggest that the proteins specifically downregulated by fluvastatin in SARS-CoV-2 infected cells may play a role in protein translation and thus SARS-CoV-2 replication.

## Discussion

Considering that a significant fraction of the population, i.e. 26% of all Americans over the age of 40 years^10^ and 30-40% of all type 2 diabetes diagnosed in Germany^24^, is on statin therapy, the primary goal of our study was to assess direct effects of statin treatment on SARS-CoV-2 infection. Our data suggest that statin treatment does not pose a risk for COVID-19 patients and fluvastatin is even associated with a mild attenuation of SARS-CoV-2 infection *in vitro*. In accordance, previous retrospectives studies reported an amelioration of COVID-19 outcome in patients^12,13,25,26^. However, given the limitations of retrospective studies, placebo-controlled trials are required to clarify the role of statins in COVID-19 infected patients. It is important to note that the inhibitory doses observed in our cell culture experiments are an order of magnitude higher than statin serum concentrations in patients, which are in the nM range^27^. This suggests that the observed direct effects on SARS-CoV-2 infection in tissue culture may not account for the clinical observations in the retrospective cohort studies. Unfortunately, fluvastatin, the only statin with anti-coronavirus activity in our study, is prescribed rarely and only one patient in the retrospective studies received this therapy^12,13^. In sum, we found no evidence for a direct aggravating effect of statins on SARS-CoV-2 infection of human lung cells.

The known immunomodulatory effects of statins^28–31^ may ameliorate the outcome of hospitalized COVID-19 patients. In our study, statin treatment did not markedly affected SARS-CoV-2 immune sensing or cytokine-mediated cellular responses in comparison to solvent control (Fig. 4). However, the slight upregulation of interferon stimulated genes (ISGs) in uninfected cells and milder dysregulation of cytokine-regulated genes after infection (Fig. 4) indicates that fluvastatin may have a relevant immunomodulatory effect on lung epithelial cells, which could ultimately ameliorate COVID-19 immunopathology. Of note, our study does not address the effect of statins on innate immune cells, which may contribute to the clinical observations. In particular, statins have a reported beneficial effect on neutrophil function in infection ^30,32,33^ and in pneumonia^34^. Investigations on the effect of statins on immune responses and neutrophil function in animal models of COVID-19 are ongoing and will clarify the clinical observations.

A possible direct inhibition of the SARS-CoV-2 main protease by statins was postulated in an *in silico* docking study^35^. In this study the binding energy for fluvastatin, simvastatin and rosuvastatin was in a similar range. Our experimental observations, however, would predict a higher efficacy and thus binding energy of fluvastatin. Hence, it is unlikely that direct targeting of the SARS-CoV-2 main protease causes the fluvastatin effects observed in this study. Alternatively, statins may indirectly inhibit SARS-CoV-2 infection by depleting cholesterol from the plasma membrane. The primary molecular target of statins is HMGCR, a rate-limiting enzyme of the mevalonate pathway that regulates *de novo* cholesterol synthesis^36^. Therefore, statins may affect SARS-CoV-2 spike - ACE2 receptor interactions. The latter was previously reported for SARS-CoV^25^. In line with this, two recent studies state that cholesterol homeostasis is critical for SARS-CoV-2 infection^37,38^. In our experimental system we only observed moderate reduction of total cellular cholesterol levels in human lung cells. However the distribution of cholesterol in the plasma membrane and hence ACE2 receptor localization may nonetheless change upon statin treatment. We are currently investigating the protein interactions of the ACE2 receptor and their cholesterol dependency to shed light on this phenomenon.

In this study, cholesterol synthesis inhibition did not cause the observed fluvastatin specific reduction of SARS-CoV-2 infection, since equimolar concentrations of simvastatin, rosuvastatin and fluvastatin affected cholesterol levels and cholesterol biosynthesis pathway enzyme expression in a comparable manner (Fig. 3, Fig. S2). This suggests that a secondary effect causes the antiviral activity of fluvastatin. Indeed, expanded target spectra of clinically approved drugs have been reported for kinase inhibitors and statins facilitating repurposing of established drugs but also aiding clinical decision making^39,40^. In line with this, fluvastatin specifically downregulated RNA binding proteins associated with protein translation and viral genome replication (Fig. 5). Some of the proteins identified in this study have been reported to modulate infection in a virus-dependent manner. For instance, PAPBC1 promotes dengue virus replication^41^ and is cleaved by HIV-1^42^ and picornaviruses^43,44^. EIF2AK2 or DDX3X are proviral proteins for the porcine reproductive and respiratory syndrome virus^45^ and hepatitis C virus^45,46^ but inhibit SARS-CoV^47^ by inducing IFN-I responses^48^. Here, we found that the eukaryotic translation initiation factor 3 subunit B (EIF3B), the RNA binding component of the eukaryotic translation factor 3 complex (EIF3), is downregulated in fluvastatin treated infected cells. As EIF3 is critical for the translation of the coronavirus murine hepatitis virus (MHV)^49^, one could speculate that EIF3B is also a host dependency factor for SARS-CoV-2. Interestingly, gene ontology annotation suggests that 80% of proteins downregulated exclusively in fluvastatin treated SARS-CoV-2 infected cells are post-translationally acetylated. Protein acetylation regulates both cellular and viral protein function, subcellular localization, and interaction^50^. Whether or not protein acetylation plays a role during SARS-CoV-2 infection requires experimental clarification.

In summary, we observed that fluvastatin downregulates host proteins required for protein translation and viral replication. Previous studies underscore the importance of these processes in SARS-CoV-2 infection^51^. Hence, we propose that fluvastatin exerts secondary effects during SARS-CoV-2 infection by reducing the abundance of SARS-CoV-2 host dependency factors. In order to pinpoint, which of the host factors involved in protein translation and viral replication are causing the observed effect, we currently verify their biological relevance during SARS-CoV-2 infection. A limitation of our study is that ACE2 was not found by tryptic fingerprinting mass spectrometry and hence we could no evaluate whether ACE2 is upregulated upon statin treatment as reported^14,15^. As we did not observe an increase in SARS-CoV-2 infection upon statin treatment, a biologically relevant ACE2 upregulation in our *in vitro* studies can be excluded. Taken together, cumulative data from this and other studies strongly argue that individuals on statin therapy can safely continue their statin regimen during the current SARS-CoV-2 pandemic.

## Methods

### Cell lines and virus strains

HEK293T cells, Huh7.5 cells constitutively expressing a Firefly luciferase (Huh7.5 FLuc) and Calu-3 cells were maintained in DMEM supplemented with 10% fetal bovine serum, 2 mM glutamine, 0.1 mM non-essential amino acids and 1% Penicillin/Streptomycin at 37°C and 5% CO_2_. Each cell line was tested regularly for Mycoplasma contamination. The recombinant hCoV229E encoding a Renilla luciferase gene was produced in Huh7.5 cells at 33°C and titrated on Huh7.5 cells. The SARS-CoV-2 isolate (strain SARS-CoV-2/München-1.2/2020/984,p3) was kindly provided by Christian Drosten (Charité, Berlin) through the European Virus Archive – Global (EVAg), amplified in Vero cells, and handled under BSL3 conditions.

### Reagents and Inhibitors

Fluvastatin (Sigma, order no. SML0038), simvastatin (Sigma, order no. S6196), atorvastatin (Sigma, order no. PZ0001), lovastatin (Sigma, order no. M2147), pravastatin (biomol, Cay-10010342) and rosuvastatin (Sigma, order no. SML1264) were dissolved in DMSO. Chloroquine (Sigma, C6628), dissolved in H_2_O, served as a positive control.

### Effect of statins on hCoV229E

Huh7.5 FLuc cells were seeded in 96-well plates at a density of 1×10^4^ cells/well in complete Dulbecco’s Modified Eagle’s Medium (DMEM) and incubated for 24 h at 37°C (5% CO_2_). The cells were pretreated for 24 h with either 5 μM of each compound or serial dilutions of the tested compounds in 100 µl final volume. Following the pretreatment, cells were inoculated with hCoV229E at MOI 0.005 in the presence of the tested compounds. The inoculum was removed 4 h later and fresh medium containing the respective compounds was added. Cells were lysed 48 h post inoculation in 50 μl detergent containing buffer and Renilla luciferase activity, as a measurement for infection, was determined by mixing 20 μl of lysate with 60 μl of luciferase substrate solution (Coelenterazine, 0.42 mg/ml in methanol). To determine cell viability, Firefly luciferase activity was measured from the same lysate by adding D-luciferin (0.2 mM D-luciferin in 25 mM glycyl-glycine) as a substrate. Luciferase activity was measured in a Centro XS3 LB960 (Berthold Technologies) microplate reader.

### SARS-2 infection and quantification by RT-qPCR

For infection experiments with SARS-CoV-2, the SARS-CoV-2 isolate (strain SARS-CoV-2/München-1.2/2020/984,p3)^18^, kindly provided by Christian Drosten (Charité, Berlin) through the European Virus Archive – Global (EVAg), was propagated in Vero cells (3 passages) after primary isolation from patient material. For infection, Calu-3 cells were seeded in collagen coated 24 well plates at a density of 4.5×10^5^ cells/well for initial infection studies or in 6 well plates at a density of 2×10^6^ cells/well for the MS data set. Cells were pretreated with 10 μM fluvastatin, simvastatin, rosuvastatin or DMSO for 24 h and then infected with SARS-CoV-2 isolate at MOI 2.0×10^−5^ based on titration in Vero cells. Heat-inactivated virus was used as a control to determine unspecific binding of virions and RNA. 4 h post infection, the inoculum was removed, cells were washed twice with PBS and fresh medium containing the respective compound was added. The infection was stopped 48 h later, viral RNA was isolated from cell lysates and supernatants using NucleoSpin RNA kit (Macherey-Nagel) and QIAamp Viral RNA Mini Kit (Qiagen), respectively, following the manufacturer’s instructions. RT-qPCR was performed in duplicates as described previously^52^ using a LightCycler 480 (Roche). In addition, SARS-CoV-2 titer in cell culture supernatants was quantified by TCID_50_ assay on Vero cells. Therefore, Vero cells were seeded in 96 well plates and incubated with serial dilutions of cell culture supernatant. 72 h later, cells were fixed with 10% formalin and stained with crystal violet.

### Cell viability assay

Cell viability was determined at the end of the experiment either by measuring Firefly luciferase activity in cell lysates (Huh7.5 Fluc cells) or by MTT assay. For the latter, MTT reagent (3-[4,5-dimethylthiazol-2-yl]-2,5 diphenyltetrazolium bromide, 0.5 mg/mL in medium) was added to the cells. The reaction was stopped with DMSO after 1 h and optical density was measured at 570 nm using a plate reader (Synergy 2, BioTek^®^).

### Primary lung cell culture

Primary human bronchial epithelial cells (HBEC) were isolated with informed consent from lung tissue from three individual patients, who underwent thoracic surgery at the University hospital of Umeå, Sweden, with ethical permission approved by the local Ethics Review Board. HBEC were grown in Bronchial Epithelial Cell Medium (BEpiCM, SC3211-b, SienCell) with recommended supplements (SC3262, SienCell) and 100 U/ml penicillin + 100 µg/ml streptomycin (PeSt; Thermo Fisher Scientific). For differentiation, 150.000 cells were seeded onto a 12 mm semipermeable transwell insert (0.4 µm Pore Polyester Membrane Insert, Corning) in differentiation media (DMEM:BEpiCM 1:1 supplemented with 52 μg/ml bovine pituitary extract, 0.5 μg/ml hydrocortisone, 0.5 ng/ml human recombinant epidermal growth factor, 0.5 μg/ml epinephrine, 10 μg/ml transferrin, 5 μg/ml insulin, 50 nM retinoic acid (all from Sigma Aldrich) and PeSt as described previously^53^. Cells were maintained submerged for the first 7 days, after which the media was removed from the apical side and cells were grown at air-liquid interface for an additional two weeks to reach full differentiation. Media was replaced trice/week with addition of fresh retinoic acid to the media shortly before usage. Differentiation of the HBEC were assessed using light microscopy focusing on epithelial morphology, presence of ciliated cells, and mucus production. Presence of ciliated cells and goblet cells was also determined with IF using antibodies directed against acetylated-tubulin and muc5AC, respectively, as described under IF staining below.

### Virus production for ALI infection

SARS-CoV-2 (SARS-CoV-2/01/human/2020/SWE; GeneBank accession no. MT093571.1) was propagated as described previously in our preprint^54^. In brief, the viral stock was propagated in Vero E6 cells for 48 h and titrated by plaque assay. For the plaque assay, 10-fold serial dilutions were added to Vero E6 cells (4*10^5^/well) seeded in 12-well plates (VWR) 12-24 h prior to infection. After 1 h incubation, the inoculum was removed, and wells were overlayed with semisolid DMEM + 2 % FBS + PeSt + 1.2 % Avicel RC/CL for incubation at 37°C in 5% CO_2_. At 65 h.p.i., the semisolid overlay was removed, and cells were fixed with 4% formaldehyde for 30 min. Cells were washed with PBS and stained with 0.5% crystal violet in 20% MeOH for 5 minutes. Plates were washed with water and the plaques counted.

### Infection and sample collection

For pretreatment at 24 h prior to infection, the medium in the basal chambers were replaced with medium containing either carrier control (DMSO), 10 µM or 50 µM fluvastatin. Immediately prior to infection, basal medium containing drug or carrier control was replaced again and the apical side of the HBEC ALI cultures was rinsed three times with PBS. For infection, 4.5×10^4^ PFU of SARS-CoV-2 (SARS-CoV-2/01/human/2020/SWE; GeneBank accession no. MT093571.1) in a total volume of 300 µl infection medium (DMEM + 10 U PeSt) was added to the apical compartment. Cells were incubated at 37°C and 5% CO_2_ for 3 h before the inoculum was removed and the cells washed one more time with PBS.

At 24, 48 and 72 h.p.i., produced and secreted virus was collected by addition of 300 µl infection medium to the apical chamber and incubation at 37°C and 5% CO_2_ for 1 h. 100 µl of the sample was used for RNA extraction using Qiaamp Viral RNA mini kit (Qiagen) according to the manufacturer’s instructions. At 48 h.p.i., the medium in the basolateral chamber was replaced with fresh medium containing drug or carrier control. After sample collection from the apical side at 72 h.p.i., the cells were washed three times with PBS and fixed over night at 4°C in 4% formaldehyde.

### Real-time (RT) q-PCR for the quantification of virus RNA in supernatant

To quantify the viral genome in the supernatant, extracted RNA was used as template in qPCRBIO Probe 1-Step Go master mix (PCR Biosystems, London, UK) together with SARS-CoV-2 primers (Forward: GTC ATG TGT GGC GGT TCA CT, Reverse: CAA CAC TAT TAG CAT AAG CAG TTG T) and probe (CAG GTG GAA CCT CAT CAG GAG ATG C) specific for viral RdRp^52^. Forward and reverse primer were modified by Magnus Lindh at Sahlgrenska University Hospital, Gothenburg. To quantify the number of viral genomes, a standard curve ranging from 2 to 2 × 103 genomes was run together with the SARS CoV-2 samples. The qPCR reaction was carried out with the StepOnePlus™ Real-Time PCR system (Thermo Fischer, Waltham, MA, USA).

### Immunofluorescence Analysis

HBEC ALI inserts were washed thrice apically and basally for 10 min with IF buffer (130 mM NaCl, 10 mM Na_2_HPO_4_/NaH_2_PO_4_, 0.05% NaN_3_, 0.1% BSA, 0.2% Triton X-100, 0.04% Tween-20, pH7.4). Inserts were blocked for 1 h at room temperature and subsequently incubated with primary antibodies (SARS: SARS-CoV-2: nucleocapsid, 40143-R001, SinoBiological; mucin5AC: Ab-1 (45M1), #MS-145-P, ThermoFisher; acetylated tubulin: T6793-0.2ml, Sigma; 1:200) overnight at 4°C in IF buffer + 10% FBS. ALI inserts were washed thrice apically and basally for 20 min with IF buffer and subsequently stained with the respective secondary antibodies (AF488: ThermoFisher A11088; AF568: ThermoFisher A21144; AF647: ThermoFisher A21240; 1:200) and Hoechst33342 (ThermoFisher 62249; 1:10000) for 1 h at room temperature in IF buffer + 10% FBS. Before imaging, samples were washed thrice apically and basally for 20 min with IF buffer and rinsed twice apically and basally for 50 min with PBS.

### Quantification

For overview pictures and quantification, whole inserts were imaged on a Cytation5 imaging platform (BioTek^®^) with installed DAPI, GFP and Cy5 LED/filter cube setups by taking 4×4 images with a 10% overlap for stitching with a 4x objective and subsequent image stitching based on the DAPI channel using linear blend fusion (Gen5 prime software, BioTek^®^). Total fluorescence intensity was quantified on a plug (region of interest) covering the maximal insert surface without touching the rim for each of the channels (Gen5 prime, BioTek^®^).

### Confocal imaging

At two sites, 4 mm punches (biopsy puncher, Miltex 33-34) were taken from the whole insert and mounted on cytoslides (Shandon, ThermoFisher 5991057) on ProLong Gold antifade mounting medium (ThermoFisher, P10144). Images were acquired on a Leica SP8 confocal microscope with a 63x oil objective at the BICU imaging unit of Umeå University. Representative sites were imaged as z-stacks of 40 slices with 0.5 µm step size and converted by maximum intensity projection using Fiji^55^.

### Mass Spectrometry

Calu-3 cells were treated with statins and infected as described in section ‘SARS-2 infection and quantification by RT-qPCR’. Cells were lysed in buffer containing 4% SDS, 10 mM DTT and 10 mM HEPES (pH 8), heated at 95°C for 10 min and sonicated at 4°C for 15 minutes (level 5, Bioruptor, Diagenode). Proteins were precipitated with acetone at −20°C and resuspended in 8 M urea buffer containing 10 mM HEPES (pH 8). Total protein was diluted to a final concentration of 100 µg/ml in 10mM HEPES (pH 8) and 2 µg total protein was used for further analysis. Reduction was performed by applying a final concentration of 5 mM TCEP and heating at 55°C for 1 h, alkylation was then performed by the addition of final concentration of 20 mM MMTS and incubation for 10 min at room temperature. Protein digestion was performed by adding trypsin at a final concentration of 1 µg protease to 50 µg total protein and incubation over night at 37°C while shaking. Peptide recovery was performed using EvoTips (Evosep) and eluted using 0.2% TFA in 60% acetonitrile (ACN). Peptides were dried and suspended in 100 µl 0.1% FA. Twenty percent of the samples were loaded on Evotips.

Samples were injected into an Evosep I HPLC (Evosep) connected to a timsTOFPro mass spectrometer with PASEF (Bruker). The 15 sample per day standard method (EvoSep I HPLC), and the standard Bruker method “PASEF method for short gradients” (MS/MS timsTOFPro) were applied.

MS/MS raw data files were processed by using Peaks X+ (Bioinformatics Solutions) and MaxQuant (1.6.17.0) software^56^ and UniProtKB databases (Homo sapiens: UP000005640/April 28, 2020; SARS-CoV-2: UP000464024/September 29, 2020). The following search parameters were used: enzyme, trypsin; maximum missed cleavages, 2; fixed modifications, methyl methanethiosulfonate (composition of H2CS at cysteine); variable modifications, oxidation (M) and acetyl (protein N-term); peptide tolerance, 20 ppm; LFQ min. ratio count, 1; Match between runs Alignment time window, 10 min; other parameters are used as default settings. All raw data files will be uploaded and made available through the Pride database. Data acquisition and analyses were performed in R (4.0.2)^57^ using following packages; tidyverse^58^, ggrepel^59^ and pheatmap^60^. All the data were transformed into log2 values and proteins with at least one valid value were used for further analysis. The missing values in each sample were replaced with a lower normal distribution (downshift = 1.8, standard deviation = 0.3). P-values from Student’s t-tests and differences of log2 values were used for the generation of volcano plots. Protein-protein interaction network analysis of statistically dysregulated proteins (p-value cut-off = 0.05) was performed with STRING with a medium confidence threshold (0.4). STRING analysis was exported to Cytoscape 3.8.2 and enrichment analysis executed with EnrichmentMap 3.1.0 plugin^61^. Pathway and upstream regulator analyses were generated through the use of Ingenuity Pathway Analysis^62^.

### Cholesterol analysis

Calu-3 cells were seeded on 24 well plates precoated with collagen. 24 h post seeding, cells were treated in duplicate with 10 µM of each statin in medium with or without FCS for two days. The cells were then washed twice in PBS, counted and stored in 1 ml HPLC-grade water. Lipid isolation was performed as previously described^63^. Samples were dissolved in methanol:acetonitrile (1:1) and quantified using a Hitachi Chromaster HPLC system fitted with a VDSpher PUR C18-H (3 μm, 150×2.0 mm) column (VDS Optilab, Berlin, Germany). The following separation conditions were used; The mobile phase consisted of methanol:acetonitire (1:1, v/v) and 0.1% formic acid dissolved in H2O (95:5), at a flow rate of 0.2 ml/min, a column temperature of 22°C and the UV detector set at 202 nm.

### Statistical analysis

Experiments were conducted in three biological replicates each with three technical replicates unless otherwise stated. All statistical analyses were performed using one or two-way analysis of variance (ANOVA) followed by Dunnett or Sidak’s multiple comparison test using GraphPad Prism software version 8 (GraphPad).

### Corresponding authors

Correspondence to Gisa Gerold

The authors declare no competing interests.

## Data Availability

No additional data reported or deposited

## Acknowledgement

G.G. was supported by the Knut and Alice Wallenberg Foundation, the Federal Ministry of Education and Research together with the Ministry of Science and Culture of Lower Saxony through the Professorinnen Programm III, and the Deutsche Forschungsgemeinschaft (DFG, German Research Foundation) - Projektnummer 158989968 - SFB 900 project C7 and the DFG project GE 2145/3-2 to G.G, the German Academic Exchange Service (DAAD) to J.K. and the Federal Ministry of Education and Research (project COVID-Protect, Projektnummer 01KI20143C) to F.J.Z.B. The work was further funded by the Infection Biology International PhD Program of Hannover Biomedical Research School to R.M., the German Centre for Infection Research (DZIF) to A.P.G. and T.P., the Helmholtz Alberta Initiative for Infectious Disease Research (HAI-IDR), the Shandong University Helmholtz International Laboratory to T.P, and the SciLifeLab/KAW national COVID-19 research program project grant 2020 and the Heart- and Lung foundation project grant for COVID-19 (project number 20200385) to A.K.Ö. and by the European Virus Archive GLOBAL (EVA-GLOBAL) project that has received funding from the European Union’s Horizon 2020 research and innovation program under grant agreement No 871029. We acknowledge the Biochemical Imaging Center at Umeå university and the National Microscopy Infrastructure, NMI (VR-RFI 2016-00968) for aiding in microscopy. We thank Anders Blomberg and Gregory Rankin at Umeå University for kindly providing us with lung tissue for isolation of HBECs. We thank Christian Drosten for providing the clinical SARS-CoV-2 isolate, Volker Thiel for the hCoV 229E strain, Sven Reiche for the SARS-CoV-2 reference material used in the RT-qPCR, Yannic Becker for assistance as well as Thomas Schulz, Tommy Olsson, Lars Nyberg and Anders Sjöstedt for constant support.

## Supplemental figures

**Fig. S1:**
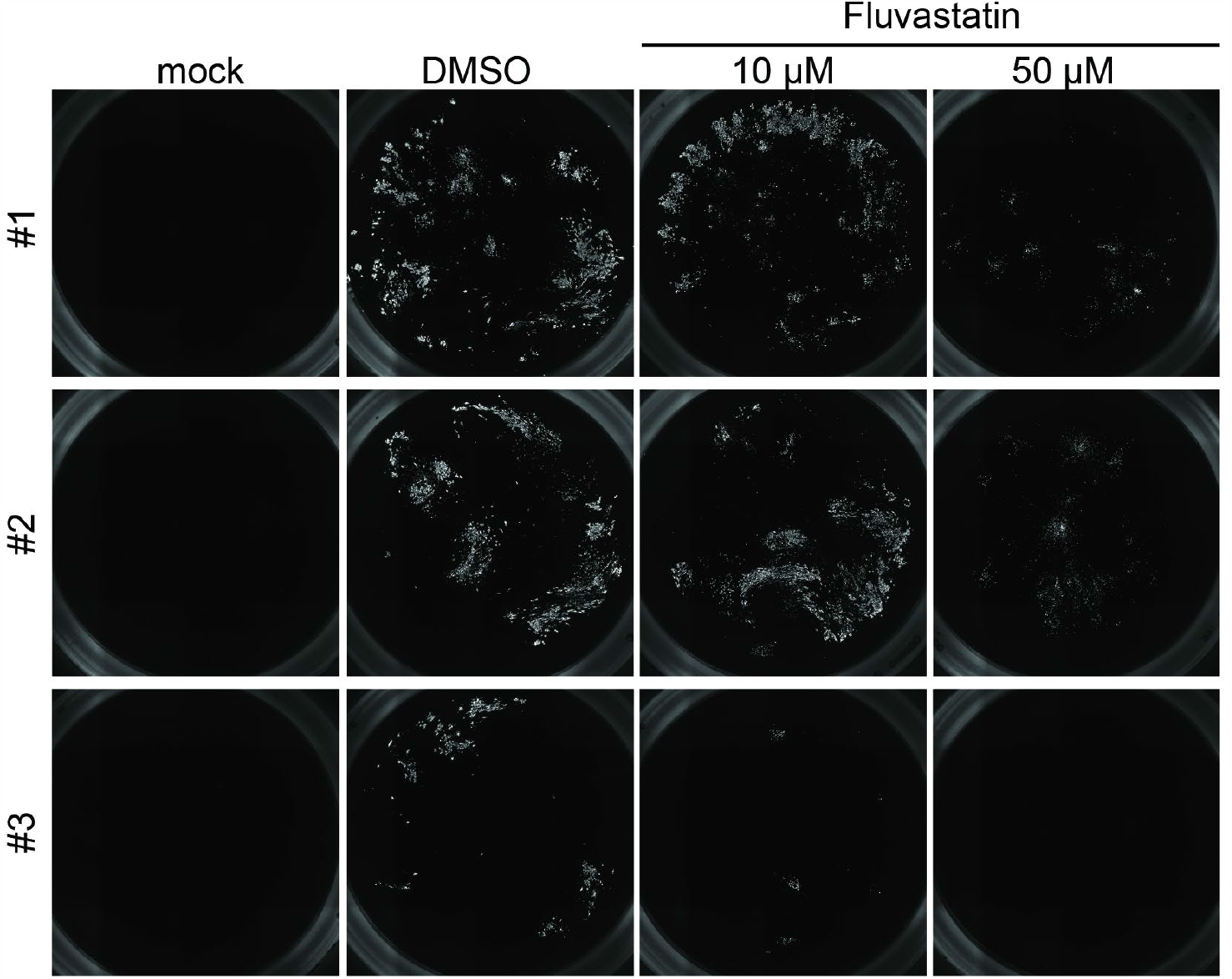
Infection of HBEC ALI cultures. HBEC ALI cultures of three individuals were infected with SARS-CoV-2, immunostained for infection (SARS-CoV-2 NC) and nuclei (Hoechst33342) and imaged by acquisition of 4×4-images per insert with a 4x object on a Cytation5 screening microscope. Depicted are representative overview images of the infection signal stitched based on the nuclear signal from each of the three donors. Stitching was performed in the BioTek Gen5+ software by alignment of images acquired with a 10% overlap using linear blend as a fusion method based on the nuclear channel and alignment was automatically transferred to the infection channel.

**Fig. S2.**
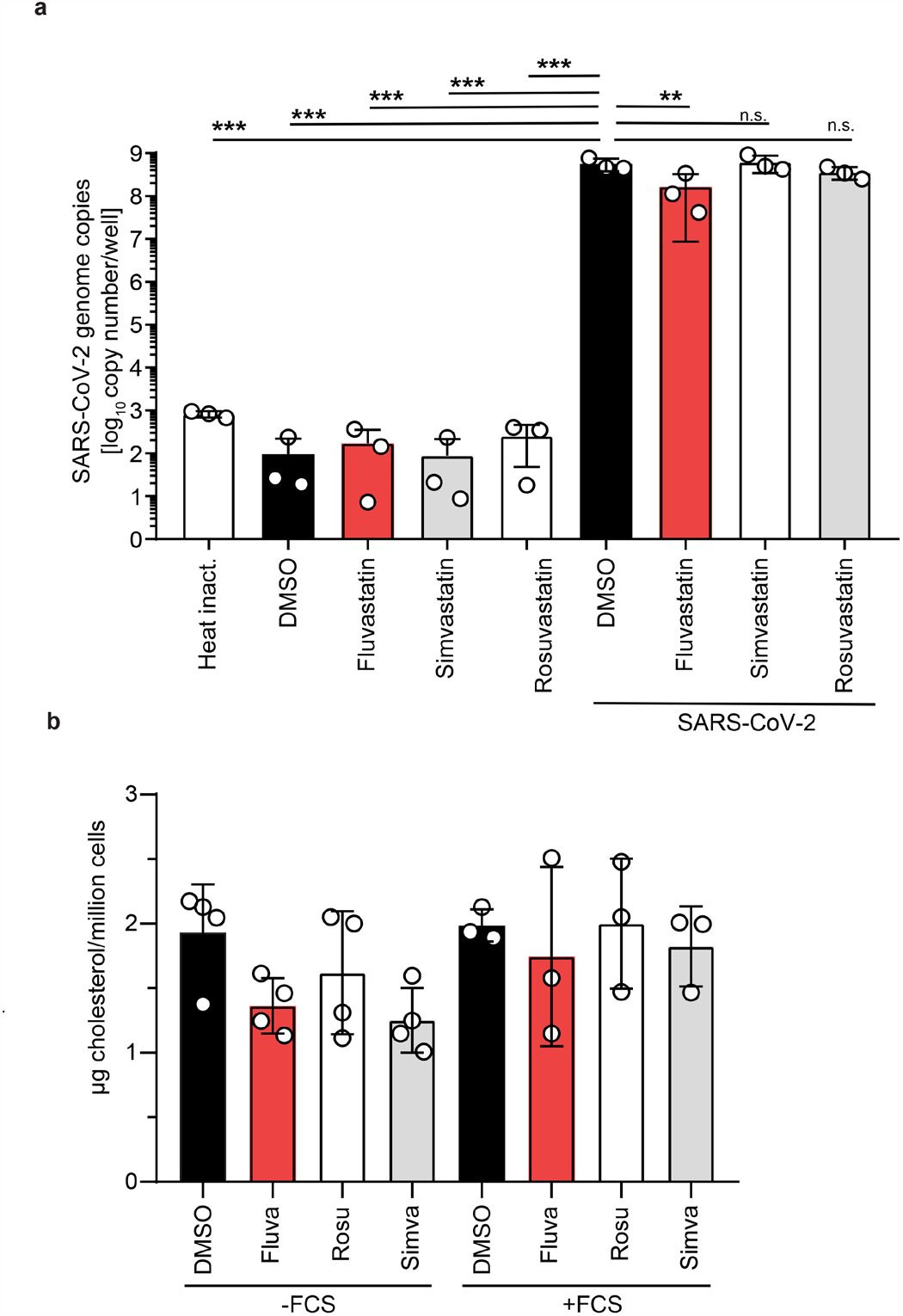
Confirmation of SARS-CoV-2 infection and effectiveness of statin treatment. **a**, Viral copy numbers in the supernatants of statin treated and SARS-CoV-2 infected Calu-3 cells that were used for the mass spectrometry analysis. Calu-3 cells were pretreated with statins or DMSO solvent control for 24 h followed by infection with SARS-CoV-2 in the presence of the drug for 48 h at an MOI of 2.×10^−5^. Viral copies were determined by RT-qPCR. n=4. **b**, Total cellular cholesterol quantification from Calu-3 cells cultured in media with or without FCS and treated with fluvastatin, rosuvastatin, simvastatin or DMSO solvent control for 3 days. Results are normalized to the corresponding DMSO solvent control. n=4 –FCS, n=3 +FCS, not significant n.s., ** P<0.01, *** P< 0.001,

**Fig. S3:**
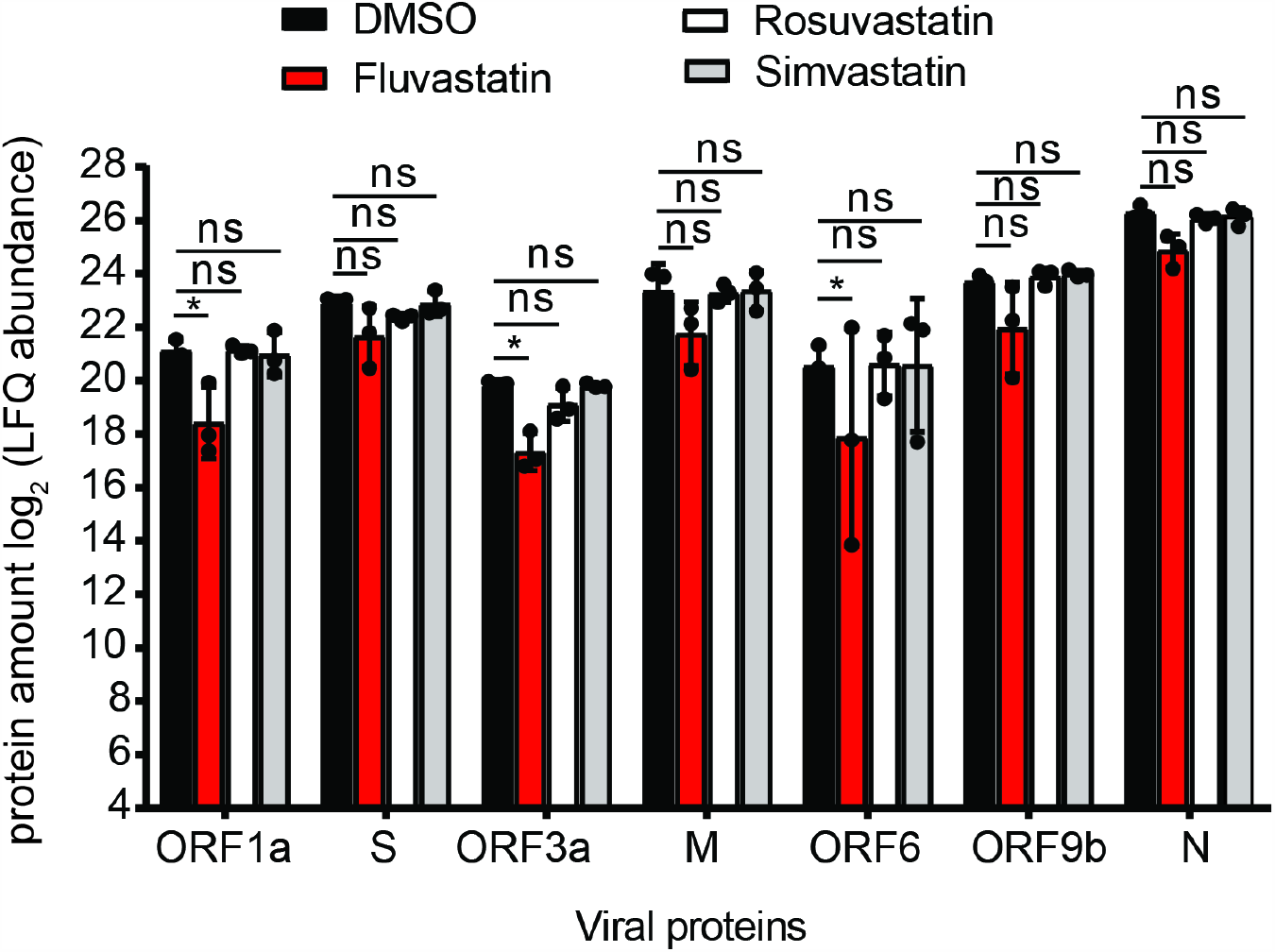
Viral protein abundance in SARS-CoV-2 infected Calu-3 cells. Abundance (as log2 values obtained by label free quantification) of seven viral proteins in SARS-CoV-2 infected Calu-3 cells treated with statins. Viral proteins were not detected in uninfected cells. For statistical analysis missing values were replaced from normal distribution with a constant downshift. One-way ANOVA, followed by Dunnett’s multiple comparison test * p<0.05, *** p<0.0005, **** p<0.0001.

**Fig. S4:**
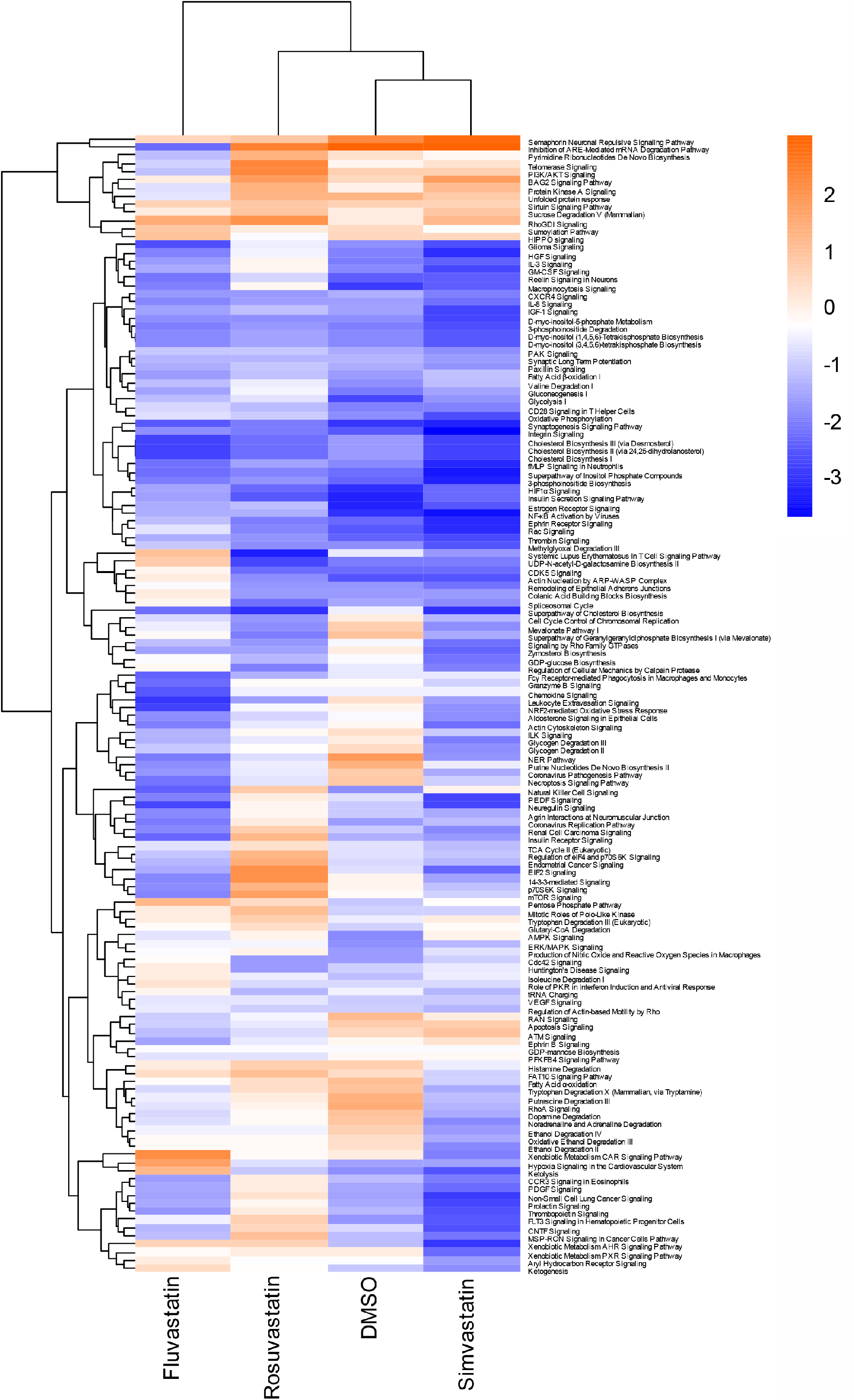
Canonical pathway analysis of SARS-CoV-2 infected Calu-3 cells. Canonical pathway analysis for the whole-cell proteome of SARS-CoV-2 infected Calu-3 cells treated with the indicated statins or DMSO solvent control. Enriched pathways with a -log10 p-value of at least ±2 in one of the three statin treatments are depicted in the heatmap. Hierarchical clustering based on enrichment score profiles. Magnification of Fig 5b.

## Notes

### Competing Interest Statement

The authors have declared no competing interest.

### Author Declarations

We have obtained ethical permission from the regionala etikprovningsnamnden in Umea to isolate human bronchial epithelial cells (HBEC) from lung resection surgery (Dnr. 2014/395-31).

### Summary of Updates

Whole cell proteomics data included

## Bibliography

1. World Health Organization. Coronavirus Disease (COVID-19) Pandemic. at <https://www.who.int/emergencies/diseases/novel-coronavirus-2019>

2. Price-Haywood, E. G., Burton, J., Fort, D. & Seoane, L. Hospitalization and Mortality among Black Patients and White Patients with Covid-19. N. Engl. J. Med. 382, 2534–2543 (2020).

3. Hoffmann, M. et al.. SARS-CoV-2 Cell Entry Depends on ACE2 and TMPRSS2 and Is Blocked by a Clinically Proven Protease Inhibitor. Cell 181, 271–280.e8 (2020).

4. Lu, Y., Liu, D. X. & Tam, J. P. Lipid rafts are involved in SARS-CoV entry into Vero E6 cells. Biochem. Biophys. Res. Commun. 369, 344–349 (2008).

5. Radenkovic, D., Chawla, S., Pirro, M., Sahebkar, A. & Banach, M. Cholesterol in Relation to COVID-19: Should We Care about It? J Clin Med 9, (2020).

6. Li, G.-M., Li, Y.-G., Yamate, M., Li, S.-M. & Ikuta, K. Lipid rafts play an important role in the early stage of severe acute respiratory syndrome-coronavirus life cycle. Microbes Infect. 9, 96–102 (2007).

7. Kozak, S. L., Heard, J. M. & Kabat, D. Segregation of CD4 and CXCR4 into distinct lipid microdomains in T lymphocytes suggests a mechanism for membrane destabilization by human immunodeficiency virus. J. Virol. 76, 1802–1815 (2002).

8. Ahn, A., Gibbons, D. L. & Kielian, M. The fusion peptide of Semliki Forest virus associates with sterol-rich membrane domains. J. Virol. 76, 3267–3275 (2002).

9. Stuart, A. D., Eustace, H. E., McKee, T. A. & Brown, T. D. K. A novel cell entry pathway for a DAF-using human enterovirus is dependent on lipid rafts. J. Virol. 76, 9307–9322 (2002).

10. Gu, Q., Paulose-Ram, R., Burt, V. L. & Kit, B. K. Prescription cholesterol-lowering medication use in adults aged 40 and over: United States, 2003-2012. NCHS Data Brief 1–8 (2014).

11. Davies, N. G. et al.. Age-dependent effects in the transmission and control of COVID-19 epidemics. Nat. Med. 26, 1205–1211 (2020).

12. Zhang, X.-J. et al.. In-Hospital Use of Statins Is Associated with a Reduced Risk of Mortality among Individuals with COVID-19. Cell Metab. 32, 176–187.e4 (2020).

13. Masana, L. et al.. Effect of statin therapy on SARS-CoV-2 infection-related. Eur. Heart J. Cardiovasc. Pharmacother. (2020). doi:10.1093/ehjcvp/pvaa128

14. Li, Y.-H. et al.. Effects of rosuvastatin on expression of angiotensin-converting enzyme 2 after vascular balloon injury in rats. J. Geriatr. Cardiol. 10, 151–158 (2013).

15. Shin, Y. H. et al.. The effect of fluvastatin on cardiac fibrosis and angiotensin-converting enzyme-2 expression in glucose-controlled diabetic rat hearts. Heart Vessels 32, 618–627 (2017).

16. Vincent, M. J. et al.. Chloroquine is a potent inhibitor of SARS coronavirus infection and spread. Virol. J. 2, 69 (2005).

17. Wang, M. et al.. Remdesivir and chloroquine effectively inhibit the recently emerged novel coronavirus (2019-nCoV) in vitro. Cell Res. 30, 269–271 (2020).

18. Wölfel, R. et al.. Virological assessment of hospitalized patients with COVID-2019. Nature 581, 465–469 (2020).

19. Ravindra, N. G. et al.. Single-cell longitudinal analysis of SARS-CoV-2 infection in human bronchial epithelial cells. BioRxiv (2020). doi:10.1101/2020.05.06.081695

20. Hui, K. P. Y. et al.. Tropism, replication competence, and innate immune responses of the coronavirus SARS-CoV-2 in human respiratory tract and conjunctiva: an analysis in ex-vivo and in-vitro cultures. Lancet Respir. Med. 8, 687–695 (2020).

21. Borthwick, F. et al.. Simvastatin treatment upregulates intestinal lipid secretion pathways in a rodent model of the metabolic syndrome. Atherosclerosis 232, 141–148 (2014).

22. Blanco-Melo, D. et al.. Imbalanced Host Response to SARS-CoV-2 Drives Development of COVID-19. Cell 181, 1036–1045.e9 (2020).

23. Allegra, A., Di Gioacchino, M., Tonacci, A., Musolino, C. & Gangemi, S. Immunopathology of SARS-CoV-2 Infection: Immune Cells and Mediators, Prognostic Factors, and Immune-Therapeutic Implications. Int. J. Mol. Sci. 21, (2020).

24. Heidemann, C., Du, Y., Baumert, J., Paprott, R. & Lampert, T. Soziale Ungleichheit und Diabetes mellitus – Zeitliche Entwicklungbei Erwachsenen in Deutschland. Journal of Health Monitoring, Robert Koch-Institut 4, (2019).

25. Glende, J. et al.. Importance of cholesterol-rich membrane microdomains in the interaction of the S protein of SARS-coronavirus with the cellular receptor angiotensin-converting enzyme 2. Virology 381, 215–221 (2008).

26. Tan, W. Y. T., Young, B. E., Lye, D. C., Chew, D. E. K. & Dalan, R. Statin use is associated with lower disease severity in COVID-19 infection. Sci. Rep. 10, 17458 (2020).

27. Björkhem-Bergman, L., Lindh, J. D. & Bergman, P. What is a relevant statin concentration in cell experiments claiming pleiotropic effects? Br. J. Clin. Pharmacol. 72, 164–165 (2011).

28. Papazian, L. et al.. Effect of statin therapy on mortality in patients with ventilator-associated pneumonia: a randomized clinical trial. JAMA 310, 1692–1700 (2013).

29. Fedson, D. S. Treating influenza with statins and other immunomodulatory agents. Antiviral Res. 99, 417–435 (2013).

30. Sapey, E. et al.. Pulmonary infections in the elderly lead to impaired neutrophil targeting, which is improved by simvastatin. Am. J. Respir. Crit. Care Med. 196, 1325–1336 (2017).

31. Pertzov, B. et al.. Hydroxymethylglutaryl-CoA reductase inhibitors (statins) for the treatment of sepsis in adults - A systematic review and meta-analysis. Clin. Microbiol. Infect. 25, 280–289 (2019).

32. Lee, C. S. et al.. Simvastatin suppresses RANTES-mediated neutrophilia in polyinosinic-polycytidylic acid-induced pneumonia. Eur. Respir. J. 41, 1147–1156 (2013).

33. Chow, O. A. et al.. Statins enhance formation of phagocyte extracellular traps. Cell Host Microbe 8, 445–454 (2010).

34. Sapey, E. et al.. Simvastatin improves neutrophil function and clinical outcomes in pneumonia. A pilot randomized controlled clinical trial. Am. J. Respir. Crit. Care Med. 200, 1282–1293 (2019).

35. Reiner, Ž. et al. Statins and the COVID-19 main protease: in silico evidence on direct interaction. Arch. Med. Sci. 16, 490–496 (2020).

36. Stancu, C. & Sima, A. Statins: mechanism of action and effects. J. Cell Mol. Med. 5, 378–387 (2001).

37. Daniloski, Z. et al.. Identification of Required Host Factors for SARS-CoV-2 Infection in Human Cells. Cell (2020). doi:10.1016/j.cell.2020.10.030

38. Wang, R. et al.. Genetic screens identify host factors for SARS-CoV-2 and common cold coronaviruses. Cell (2020). doi:10.1016/j.cell.2020.12.004

39. Huang, Y. et al.. A framework for identification of on- and off-target transcriptional responses to drug treatment. Sci. Rep. 9, 17603 (2019).

40. Klaeger, S. et al.. The target landscape of clinical kinase drugs. Science 358, (2017).

41. Suzuki, Y. et al.. Characterization of RyDEN (C19orf66) as an Interferon-Stimulated Cellular Inhibitor against Dengue Virus Replication. PLoS Pathog. 12, e1005357 (2016).

42. Alvarez, E., Castelló, A., Menéndez-Arias, L. & Carrasco, L. HIV protease cleaves poly(A)-binding protein. Biochem. J. 396, 219–226 (2006).

43. Rivera, C. I. & Lloyd, R. E. Modulation of enteroviral proteinase cleavage of poly(A)-binding protein (PABP) by conformation and PABP-associated factors. Virology 375, 59–72 (2008).

44. Zhang, B., Morace, G., Gauss-Müller, V. & Kusov, Y. Poly(A) binding protein, C-terminally truncated by the hepatitis A virus proteinase 3C, inhibits viral translation. Nucleic Acids Res. 35, 5975–5984 (2007).

45. Wang, X., Zhang, H., Abel, A. M. & Nelson, E. Protein kinase R (PKR) plays a pro-viral role in porcine reproductive and respiratory syndrome virus (PRRSV) replication by modulating viral gene transcription. Arch. Virol. 161, 327–333 (2016).

46. Li, Q., Pène, V., Krishnamurthy, S., Cha, H. & Liang, T. J. Hepatitis C virus infection activates an innate pathway involving IKK-α in lipogenesis and viral assembly. Nat. Med. 19, 722–729 (2013).

47. de Wilde, A. H. et al.. A Kinome-Wide Small Interfering RNA Screen Identifies Proviral and Antiviral Host Factors in Severe Acute Respiratory Syndrome Coronavirus Replication, Including Double-Stranded RNA-Activated Protein Kinase and Early Secretory Pathway Proteins. J. Virol. 89, 8318–8333 (2015).

48. Lai, M.-C., Sun, H. S., Wang, S.-W. & Tarn, W.-Y. DDX3 functions in antiviral innate immunity through translational control of PACT. FEBS J. 283, 88–101 (2016).

49. V’kovski, P. et al. Determination of host proteins composing the microenvironment of coronavirus replicase complexes by proximity-labeling. Elife 8, (2019).

50. Murray, L. A., Combs, A. N., Rekapalli, P. & Cristea, I. M. Methods for characterizing protein acetylation during viral infection. Meth. Enzymol. 626, 587–620 (2019).

51. Bojkova, D. et al. Proteomics of SARS-CoV-2-infected host cells reveals therapy targets. Nature 583, 469–472 (2020).

52. Corman, V. M. et al. Detection of 2019 novel coronavirus (2019-nCoV) by real-time RT-PCR. Euro Surveill. 25, (2020).

53. Danahay, H., Atherton, H., Jones, G., Bridges, R. J. & Poll, C. T. Interleukin-13 induces a hypersecretory ion transport phenotype in human bronchial epithelial cells. Am. J. Physiol. Lung Cell Mol. Physiol. 282, L226–36 (2002).

54. Rosendal, E. et al. Detection of asymptomatic SARS-CoV-2 exposed individuals by a sensitive S-based ELISA. medRxiv (2020). doi:10.1101/2020.06.02.20120477

55. Schindelin, J. et al. Fiji: an open-source platform for biological-image analysis. Nat. Methods 9, 676–682 (2012).

56. Tyanova, S., Temu, T. & Cox, J. The MaxQuant computational platform for mass spectrometry-based shotgun proteomics. Nat. Protoc. 11, 2301– 2319 (2016).

57. R Core Team (2020). R: A language and environment for statistical computing. R Foundation for Statistical Computing, Vienna, Austria. at <https://www.R-project.org/>

58. Wickham, H. et al. Welcome to the tidyverse. JOSS 4, 1686 (2019).

59. Kamil Slowikowski. ggrepel: Automatically PositionNon-Overlapping Text Labels with “ggplot2”. R package version 0.8.2. (2020). at <https://CRAN.R-project.org/package=ggrepel>

60. Raivo Kolde. pheatmap:Pretty Heatmaps. Rpackage version 1.0.12. (2019). at <https://CRAN.R-project.org/package=pheatmap>

61. Merico, D., Isserlin, R., Stueker, O., Emili, A. & Bader, G. D. Enrichment map: a network-based method for gene-set enrichment visualization and interpretation. PLoS One 5, e13984 (2010).

62. Krämer, A., Green, J., Pollard, J. & Tugendreich, S. Causal analysis approaches in Ingenuity Pathway Analysis. Bioinformatics 30, 523–530 (2014).

63. Beck, A. et al. Quantification of sterols from carp cell lines by using HPLC-MS. Sep. Sci. plus 1, 11– 21 (2018).

